## Supplementary material for "Implementation strategies to increase human papillomavirus vaccination uptake for adolescent girls in sub-Saharan Africa: A scoping review protocol": S1 Appendix 1 PubMed search

**S1_Appendix 1 PUBMED SEARCH 26.01.2022.**

| **Search** | **Actions** | **Details** | **Query** | **Results** | **Time** |
| --- | --- | --- | --- | --- | --- |
| #6 |  |  | Search: **((((#1) AND (#2)) AND (#3)) AND (#4)) AND (#5)** | [142](https://pubmed.ncbi.nlm.nih.gov/?term=%28%28%28%28%231%29+AND+%28%232%29%29+AND+%28%233%29%29+AND+%28%234%29%29+AND+%28%235%29&sort=) | 07:38:37 |
| #5 |  |  | Search: **((((((((((((((((((((((((((((((((((((((((((((((((sub-Saharan Africa) OR (Africa south Sahara)) OR (SSA)) OR (Angola)) OR (Benin)) OR (Burkina Faso)) OR (Botswana)) OR (Burundi)) OR (Cameroon)) OR (Cape Verde)) OR (Central Africa Republic)) OR (Chad)) OR (Congo)) OR (Cote d'voire)) OR (Djibouti)) OR (Eritrea)) OR (Ethiopia)) OR (The Gambia)) OR (Ghana)) OR (Guinea)) OR (Guinea-Bissau)) OR (Gabon)) OR (Kenya)) OR (Lesotho)) OR (Liberia)) OR (Madagascar)) OR (Malawi)) OR (Mali)) OR (Mauritania)) OR (Mauritius)) OR (Mozambique)) OR (Namibia)) OR (Niger)) OR (Nigeria)) OR (Rwanda)) OR (Sao tome principle)) OR (Senegal)) OR (seychelles)) OR (sierra Leon)) OR (Somalia)) OR (South Africa)) OR (Sudan)) OR (south Sudan)) OR (Swaziland)) OR (United republic of Tanzania)) OR (Togo)) OR (Uganda)) OR (Zimbabwe)) OR (Zambia)** | [635,885](https://pubmed.ncbi.nlm.nih.gov/?term=%28%28%28%28%28%28%28%28%28%28%28%28%28%28%28%28%28%28%28%28%28%28%28%28%28%28%28%28%28%28%28%28%28%28%28%28%28%28%28%28%28%28%28%28%28%28%28%28sub-Saharan+Africa%29+OR+%28Africa+south+Sahara%29%29+OR+%28SSA%29%29+OR+%28Angola%29%29+OR+%28Benin%29%29+OR+%28Burkina+Faso%29%29+OR+%28Botswana%29%29+OR+%28Burundi%29%29+OR+%28Cameroon%29%29+OR+%28Cape+Verde%29%29+OR+%28Central+Africa+Republic%29%29+OR+%28Chad%29%29+OR+%28Congo%29%29+OR+%28Cote+d%27voire%29%29+OR+%28Djibouti%29%29+OR+%28Eritrea%29%29+OR+%28Ethiopia%29%29+OR+%28The+Gambia%29%29+OR+%28Ghana%29%29+OR+%28Guinea%29%29+OR+%28Guinea-Bissau%29%29+OR+%28Gabon%29%29+OR+%28Kenya%29%29+OR+%28Lesotho%29%29+OR+%28Liberia%29%29+OR+%28Madagascar%29%29+OR+%28Malawi%29%29+OR+%28Mali%29%29+OR+%28Mauritania%29%29+OR+%28Mauritius%29%29+OR+%28Mozambique%29%29+OR+%28Namibia%29%29+OR+%28Niger%29%29+OR+%28Nigeria%29%29+OR+%28Rwanda%29%29+OR+%28Sao+tome+principle%29%29+OR+%28Senegal%29%29+OR+%28seychelles%29%29+OR+%28sierra+Leon%29%29+OR+%28Somalia%29%29+OR+%28South+Africa%29%29+OR+%28Sudan%29%29+OR+%28south+Sudan%29%29+OR+%28Swaziland%29%29+OR+%28United+republic+of+Tanzania%29%29+OR+%28Togo%29%29+OR+%28Uganda%29%29+OR+%28Zimbabwe%29%29+OR+%28Zambia%29&sort=) | 07:37:13 |
| #4 |  |  | Search: **(Uptake) OR (coverage)** | [565,814](https://pubmed.ncbi.nlm.nih.gov/?term=%28Uptake%29+OR+%28coverage%29&sort=) | 07:35:27 |
| #3 |  |  | Search: **((Human papillomavirus vaccination) OR (HPV vaccination)) OR (HPV immunisation)** | [18,234](https://pubmed.ncbi.nlm.nih.gov/?term=%28%28Human+papillomavirus+vaccination%29+OR+%28HPV+vaccination%29%29+OR+%28HPV+immunisation%29&sort=) | 07:34:36 |
| #2 |  |  | Search: **(Implementation strategies) OR (interventions)** | [9,425,631](https://pubmed.ncbi.nlm.nih.gov/?term=%28Implementation+strategies%29+OR+%28interventions%29&sort=) | 07:33:47 |
| #1 |  |  | Search: **((((((Girls) OR (parents)) OR (Teachers)) OR (Health care providers)) OR (health professionals)) OR (doctors)) OR (nurses)** | [3,168,662](https://pubmed.ncbi.nlm.nih.gov/?term=%28%28%28%28%28%28Girls%29+OR+%28parents%29%29+OR+%28Teachers%29%29+OR+%28Health+care+providers%29%29+OR+%28health+professionals%29%29+OR+%28doctors%29%29+OR+%28nurses%29&sort=) | 07:31:21 |
