## Supplementary material for "Implementation strategies to increase human papillomavirus vaccination uptake for adolescent girls in sub-Saharan Africa: A scoping review protocol": S2 Appendix 2 data extraction form

| First Author full names, publication year, country | Year of study | Title | Type of programme | Study design* | Sample size | Targeted stakeholder | Frame work | Age of girls/school grade | Funding source | Data collecting tool | Implementation strategies | HPV vaccine coverage | HPV vaccine uptake |
| --- | --- | --- | --- | --- | --- | --- | --- | --- | --- | --- | --- | --- | --- |

**SAMPLE DATA CHARTING FORM**
