## Supplementary material for "Implementation strategies to increase human papillomavirus vaccination uptake for adolescent girls in sub-Saharan Africa: A scoping review protocol": S3 Appendix 3 PRISMA ScR Checklist

**S3 Checklist: Preferred Reporting Items for Systematic reviews and Meta-Analyses extension for Scoping Reviews (PRISMA-ScR) Checklist**

| **SECTION** | **ITEM** | **PRISMA-ScR CHECKLIST ITEM** | **REPORTED ON PAGE #** |
| --- | --- | --- | --- |
| **TITLE** | | | |
| Title | 1 | Implementation strategies to increase human papillomavirus vaccination uptake for adolescent girls in sub-Saharan Africa: A scoping review protocol | 1 |
| **ABSTRACT** | | | |
| Structured summary | 2 | Abstract provided | 1 |
| **INTRODUCTION** | | | |
| Rationale | 3 | This review will provide a basis for policy on increasing uptake of HPV vaccine for adolescent girls and future research including effectiveness systematic reviews | 4 |
| Objectives | 4 | **Review question**  What implementation strategies have increased the uptake of HPV vaccination for adolescent girls in SSA?  **Objective:** To identify implementation strategies to increase HPV vaccination uptake for adolescent girls in sub-Saharan Africa and provide a basis for future systematic reviews to evaluate effective strategies. | 5 |
| **METHODS** | | | |
| Protocol and registration | 5 | Protocol registration https://osf.io/gfd7s/. | 5 |
| Eligibility criteria | 6 | We intend to capture all research papers published in English on implementation strategies including various stakeholders (parents, adolescent girls, teachers and community leaders between January 2006 and December 2021 | 5 |
| Information sources* | 7 | The databases to be searched include MEDLINE (via PubMed), EMBASE, CINAHL (Cumulative Index to Nursing and Allied Health Literature) (EBSCO) and Scopus. Grey literature citations will also be searched including Google scholar | 5 |
| Search | 8 | Provided as S1 | S1 Appendix 1 |
| Selection of sources of evidence† | 9 | Two independent reviewers will assess articles for inclusion by screening titles, abstracts ad full text | 7 |
| Data charting process‡ | 10 | The data extracted will include specific details about the participants, concept, context, study methods and key findings relevant to the review question. The draft charting tool will be piloted on 5 included pieces of evidence and revisions will be made as necessary during the process of data charting from each included evidence source. Modifications will be detailed in the scoping review. Authors of papers will be contacted to request missing or additional data, where required, if no response is received, available data will be used in the best possible way. Data charting will be done by two independent reviewers. | 7 |
| Data items | 11 | Also provided as S2  1. First author  2. Year of study  3. Year of publication  4. Country(ies)  5. Title  6. Type of program  7. Study design (where applicable)  8. Sample size  9. Targeted stakeholders  10. Framework  11. Age of girls/ school grade  12. Implementation strategies  13. Number of implementation strategies  14. Funding source  15. HPV vaccine uptake  16. HPV vaccine coverage | 8 |
| Critical appraisal of individual sources of evidence§ | 12 | NA | NA |
| Synthesis of results | 13 | The extracted data will be presented in table or graphical forms to align with the study objectives. These will be accompanied by a narrative summary of how the findings relate to the research question and objectives. | 8 |
| **RESULTS** | | | |
| Selection of sources of evidence | 14 |  | Click here to enter text. |
| Characteristics of sources of evidence | 15 |  | Click here to enter text. |
| Critical appraisal within sources of evidence | 16 |  | Click here to enter text. |
| Results of individual sources of evidence | 17 |  | Click here to enter text. |
| Synthesis of results | 18 |  | Click here to enter text. |
| **DISCUSSION** | | | |
| Summary of evidence | 19 | **Objective:** To identify implementation strategies to increase HPV vaccination uptake for adolescent girls in sub-Saharan Africa and provide a basis for future systematic reviews to evaluate effective strategies. | 8 |
| Limitations | 20 | This scoping review will only focus on studies done in English hence may miss other important research done in other languages, further only strategies focusing on uptake of the vaccine for girls only will be considered, yet there could be other strategies used for vaccine uptake among boys | 8 |
| Conclusions | 21 |  | Click here to enter text. |
| **FUNDING** | | | |
| Funding | 22 |  | Click here to enter text. |

JBI = Joanna Briggs Institute; PRISMA-ScR = Preferred Reporting Items for Systematic reviews and Meta-Analyses extension for Scoping Reviews.

* Where *sources of evidence* (see second footnote) are compiled from, such as bibliographic databases, social media platforms, and Web sites.

† A more inclusive/heterogeneous term used to account for the different types of evidence or data sources (e.g., quantitative and/or qualitative research, expert opinion, and policy documents) that may be eligible in a scoping review as opposed to only studies. This is not to be confused with *information sources* (see first footnote).

‡ The frameworks by Arksey and O’Malley (6) and Levac and colleagues (7) and the JBI guidance (4, 5) refer to the process of data extraction in a scoping review as data charting*.*

§ The process of systematically examining research evidence to assess its validity, results, and relevance before using it to inform a decision. This term is used for items 12 and 19 instead of "risk of bias" (which is more applicable to systematic reviews of interventions) to include and acknowledge the various sources of evidence that may be used in a scoping review (e.g., quantitative and/or qualitative research, expert opinion, and policy document).

*From:* Tricco AC, Lillie E, Zarin W, O'Brien KK, Colquhoun H, Levac D, et al. PRISMA Extension for Scoping Reviews (PRISMAScR): Checklist and Explanation. Ann Intern Med. 2018;169:467–473. [doi: 10.7326/M18-0850](http://annals.org/aim/fullarticle/2700389/prisma-extension-scoping-reviews-prisma-scr-checklist-explanation).
